# Newborn dried blood spots for serological surveys of COVID-19

**DOI:** 10.1101/2020.08.14.20175299

**Authors:** Feimei Liu, Mytien Nguyen, Pavithra Vijayakumar, Alanna Kaplan, Amit Meir, Yile Dai, Eric Wang, Hannah Walsh, Aaron M. Ring, Saad B. Omer, Shelli F. Farhadian

## Abstract

There is an urgent need for inexpensive, population-wide surveillance testing for COVID-19. We tested newborn dried blood spots (DBS) anti-SARS-COV-2 antibodies for all infants born at Yale from March-May 2020, and found that newborn DBS serologies reflect maternal and population-wide infection rates during the study period. This suggests a role for DBS in COVID-19 surveillance in areas where viral testing is limited.

## Introduction

Population seroprevalence surveys are important tools for COVID-19 public health decision-making. However, there are significant hurdles to implementing large-scale serological surveys, especially in resource poor settings. Newborn dried blood spot (DBS) is an easy, inexpensive, and widely used method to collect and store newborn blood specimens, and, in cases where newborn antibody results reflect maternal seropositivity, DBS is an appealing tool to measure COVID-19 population seroprevalence trends among pregnant women.^1^ Here, we evaluated DBS as a potential COVID-19 surveillance tool.

## Methods

Discarded newborn DBS was obtained for all infants born in the Yale New Haven hospital system from February 19-May 26, 2020, a time period that began two weeks prior to the first confirmed case in Connecticut (March 8). Routine SARS-CoV-2 PCR testing of all women who presented to labor and delivery began on April 1, 2020. We adapted a standardized ELISA Serology Assay^2^ for detecting anti-SARS-CoV-2 antibody using newborn DBS. Anti-SARS-CoV-2 IgG was measured in all samples, and IgM was measured where the IgG result was positive. The threshold for a positive result was set at the 99% confidence interval, at which 1% of true negatives would be reported as false positives. Banked cord blood from infants born prior to 2019 was run as negative controls. Maternal demographic and clinical features were examined in association with the likelihood of an antibody positive newborn using multivariable logistic regression performed in STATA 16.1.

This study was deemed exempt from human subjects regulations by the Yale institutional review board.

## Results

The first positive DBS in the study was observed from an infant born in Connecticut on February 18, 2020 **(Figure 1)**. Overall 182/3048 (5.9%) DBS tested were positive for anti-SARS-CoV-2 IgG. None were positive for anti-SARS-CoV-2 IgM. Of the 182 infants with positive antibody results, 134 had mothers who underwent testing for COVID-19 by viral PCR prior to delivery, of which 65 (49%) had a positive test and 69 (51%) had a negative PCR test. All pre-2019 cord blood samples were negative for SARS-CoV-2 IgG by this assay.

**Figure 1:**
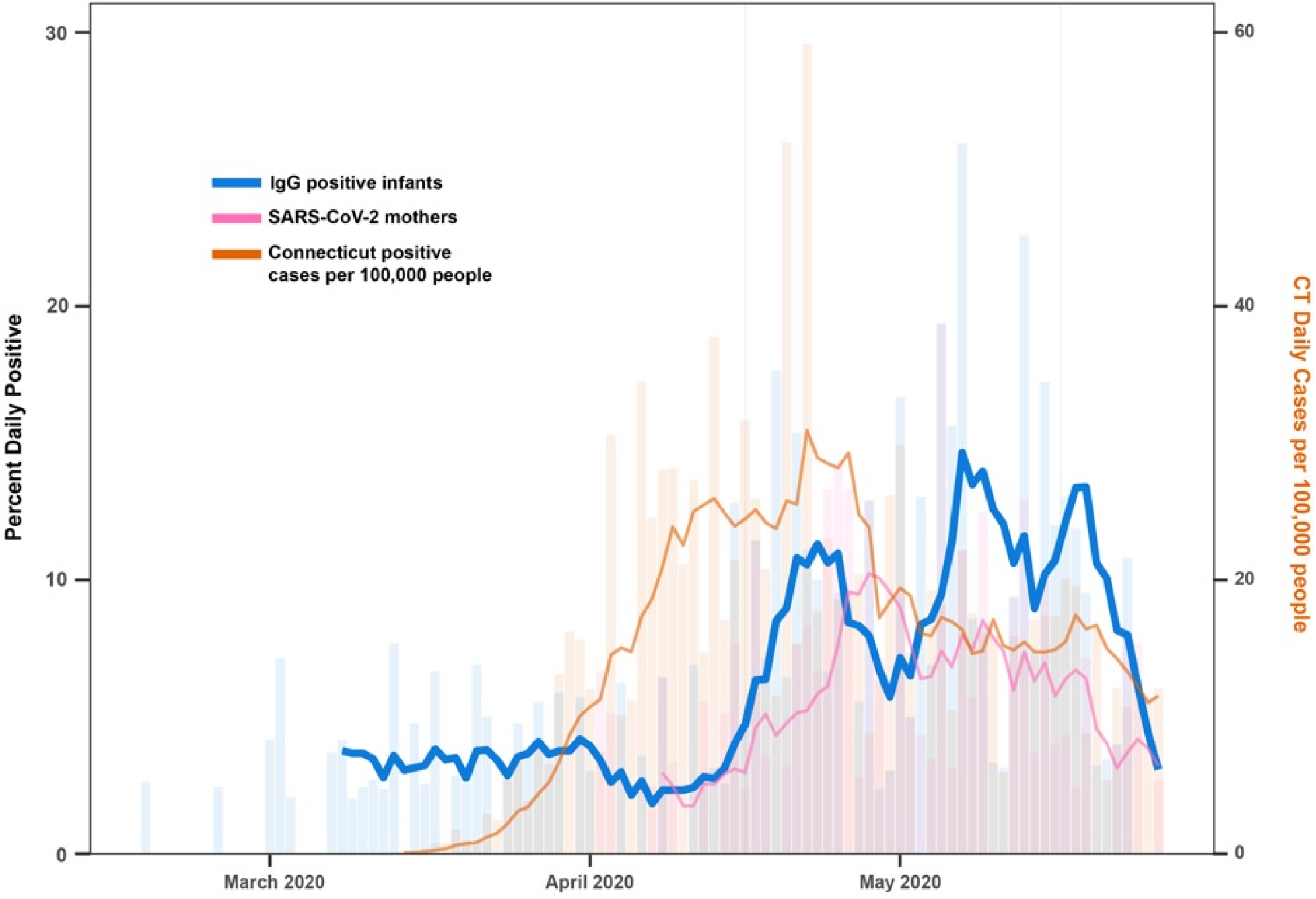
Rate of newborn COVID-19 positive antibody and cases in Connecticut from February to May 2020. Daily positive rate (bars) and 7-day moving average (lines) of newborn IgG antibody (blue) in relation to SARS-CoV-2 PCR in mothers (pink), and positive cases per 100,000 people in Connecticut State (orange). Connecticut statewide data were retrieved from Hughes *et al*. (2020) and rate per 100,000 calculated using U.S. Census estimates.

We examined the inter-relationship between daily newborn IgG antibody positive rates to maternal and statewide COVID-19 infection rates. The daily positive proportion of DBS specimens were predicted strongly by both maternal SARS-CoV-2 infection rate (*p*<0.001) and statewide COVID-19 daily positive test counts per 100,000 people (*p*=0.010). Further investigations using cross-correlation analysis demonstrated that daily newborn IgG antibody positive rate strongly correlated with maternal SARS-CoV-2 infection rate at zero lag time (*r*=0.47), and statewide COVID-19 positive test counts at a lag-time of 15-days (*r*=0.45).

We examined maternal demographics and clinical factors that may be associated with the likelihood of an lgG+ newborn. Of the 92 mothers who tested positive for SARS-CoV-2 during the universal screening period, 90 had complete demographic and clinical data. When controlled for body mass index (BMI), hypertension, diabetes, COVID-19 symptoms, gestational age, and time between maternal COVID-19 diagnosis and delivery (lag time), infants that were antibody positive for COVID-19 were more likely to be born later during the study period (adjusted OR, 1.05; 95% CI, 1.01-1.10, *p*=0.01), and to mothers with older maternal age (adjusted OR, 1.13; 95% CI, 1.02-1.25, *p*=0.01). No significant associations were found for the other factors.

## Discussion

Our findings demonstrate utility of the newborn DBS SARS-CoV-2 antibody assay to detect past maternal infection and suggest a use for DBS to measure population-level trends of COVID-19, as well as a way to monitor for resurgence of this disease. The detection of seropositive newborns prior to the availability of viral testing in CT indicates that DBS surveillance may be a useful tool for COVID-19 surveillance where viral testing is limited. Most, but not all, mothers who screened positive for SARS-CoV-2 during the study period delivered a newborn with detectable anti-SAR-CoV-2 IgG antibody, a finding that may reflect the lag time in development of detectable antibodies after infection.^3^

This study has limitations. Most mothers were screened for SARS-CoV-2 at the time of hospitalization for delivery, but we were unable to determine the true date of maternal infection. Because we did not have access to the date of infection or serum antibody testing results for all mothers, we could not determine whether mothers who screened positive for SARS-CoV-2 by PCR but delivered a seronegative newborn had poor antibody responses themselves, had a remote infection with antibody responded that waned prior to delivery, or whether there was inefficient transplacental transfer of antibody in these cases. However, given that the majority of PCR positive mothers delivered seropositive newborns, this does not appear to diminish the utility of DBS as a surveillance tool.

In this study, we demonstrate that levels of IgG in DBS reflect overall population-level trends in case incidence, with a lag that is consistent with the time to development of detectable antibodies after infection, making DBS antibody testing an attractive option for large-scale population surveillance during the COVID-19 pandemic. As dried blood spots are routinely collected from newborns, no additional sample collection is required, and specimens can be stored for later assessment. Using DBS as a surveillance tool may therefore be particularly advantageous in resource-poor settings, where innovative tools of field epidemiology will be required in order to control the spread of the virus.

## Data Availability

All data stored on HIPAA-secure server; access may be available upon request

## Author Contributions

SFF had full access to all of the data in the study and takes responsibility for the integrity of the data and the accuracy of the data analysis.

Concept and design: SFF and SBO

Acquisition, analysis, or interpretation of data: All authors.

Drafting of the manuscript: SFF, MN, PV, FL

Critical revision of the manuscript for important intellectual content: All authors.

Statistical analysis: MN, PV, FL

Administrative, technical, or material support: AR, SFF

Supervision: SFF

## Additional contributions

We acknowledge Dr. Wade Schulz who assisted with data acquisition.

## References

1. Bjorkesten J, Enroth S, Shen Q, et al. Stability of Proteins in Dried Blood Spot Biobanks. Mol Cell Proteomics. 2017;16(7):1286-1296.

2. Amanat F, Stadlbauer D, Strohmeier S, et al. A serological assay to detect SARS-CoV-2 seroconversion in humans. Nat Med. 2020.

3. Long QX, Liu BZ, Deng HJ, et al. Antibody responses to SARS-CoV-2 in patients with COVID-19. Nature medicine. 2020;26(6):845-848

